# ‘I Gotta Feelee-ing’: Exploring the Effects of a Smartphone app (Feelee) to Enhance Adolescents’ Emotion Regulation in Forensic Outpatient Settings. A Multiple Single-Case Experimental Design

**DOI:** 10.1101/2025.09.08.25334620

**Authors:** Merel M.L. Leijse, Levi van Dam, Samantha Bouwmeester, Thimo M. van der Pol, René Breuk, Arne Popma

## Abstract

Adolescents in forensic outpatient care often face a complex interplay of emotional and cognitive challenges, frequently shaped by adverse childhood experiences, increasing their risk for delinquent behavior. Current interventions show mixed results, potentially due to a mismatch between intervention demands and adolescents disrupted emotional development. The Feelee app may help bridge this gap by offering daily practice of emotion regulation skills through the collection of active and passive smartphone data. This study aimed to assess the initial effectiveness of the Feelee app as an addition to treatment as usual to enhance emotion regulation skills among forensic outpatients. A multiple single-case experimental ABA design was applied over a 2-week baseline (phase A^1^), 4-week intervention (phase B), and 2-week follow-up (phase A^2^), combining quantitative and qualitative methods. Twenty-two adolescents (aged 12–23) completed daily assessments of emotion regulation. Secondary outcomes focused on emotional developmental mechanisms and treatment-related factors, measured at pre-, post-, and follow-up. Qualitatively, semi-structured interviews with adolescents and clinicians explored experiences with Feelee and its integration into treatment. Results showed a significant reduction in emotional suppression during the intervention phase. No improvements were found in emotion recognition and impulse control, while reflection and distraction showed reversed outcomes. On secondary outcomes, the follow-up measurements revealed increased positive emotion differentiation, emotional awareness and self-reflection. Treatment motivation remained stable, while therapeutic alliance improved. Qualitative findings highlighted increased emotional insight but also pointed to technical difficulties and limited discussion of Feelee data during sessions. These results suggest that Feelee may be particularly helpful in the early stages of emotion regulation by reducing suppression, making it possible to take the crucial first step of engaging with emotions. Future research should explore longer-term use and actively involve clinicians in the integration of app data to maximize therapeutic relevance and impact.

## Introduction

Adolescents receiving forensic outpatient care often face a complex interplay of psychological, social and behavioral challenges that are associated with the risk for delinquent behavior [1–3]. These multiproblem behaviors are strongly linked to adverse childhood experiences (ACEs), such as parental neglect, maltreatment and poverty [4–6]. Such traumatic experiences increase the likelihood of additional risk factors, including family conflicts, poor school performances and bad peer influences, which are well-known contributors to delinquent behavior [7–10]. Moreover, the accumulation of life stressors and risk factors has been shown to impair both emotional and cognitive development, reducing adolescents’ capacity for effective emotion regulation and insight into problematic behavior [8,11–13]. These multi problems in adolescence highlight the complexity and need for effective, tailored interventions for adolescents at risk for delinquent behavior.

Current interventions in forensic outpatient settings primary aims to address the multiproblem by focusing on reducing risk factors and increasing protective factors. While these approaches are considered evidence-based, their effectiveness remains mixed [14,15]. Meta-analyses report small to moderate effects for family-based interventions and inconsistent outcomes for individually oriented CBT in reducing delinquent behavior [16–18]. These inconclusive results may be attributed to methodological limitations, but more fundamentally may reflect a mismatch between the demands of these interventions and the developmental and psychosocial profiles of the adolescents involved. Many adolescents in forensic care face disrupted emotional and cognitive development, limiting their emotional insight, capacity for reflection and regulation which are skills that are often presumed in standard therapeutic approaches [19–21]. This limited insight into their own emotions and behavior not only hampers emotional learning but also reduces engagement and intrinsic motivation in treatment [22–24]. Adolescents report they do not feel involved or represented in the goals and methods of treatment, which reduces their motivation and diminishes the potential impact of the intervention [25]. As a result, current interventions may fall short of addressing the actual needs and capabilities of adolescents in forensic outpatient care.

Nonetheless, in recent years, there has been an increasing interest in the use of technologies to enhance traditional treatments in more personalized and continuous (24/7) interventions [26–28]. Smartphones, in particular, offer accessible functionalities that enable users to easily access therapeutic content and engage in skill practice in real time and naturalistic settings [29,30]. Additionally, smartphones facilitate the collection of both active and passive data [31,32]. Active data are generated through intentional user input, such as ecological momentary assessments/experience sampling methods (EMA/ESM). Passive data, in contrast, are automatically gathered through embedded sensors like GPS, accelerometers, and usage patterns. Together, these data streams contribute to the creation of a digital phenotype, of which the self-report and objectively measured data provide valuable insights into in emotion and behavioral patterns [33,34].

Feelee is a smartphone application that integrates both active and passive data sources to support emotion regulation skills in adolescence. Active data are gathered through digital diary entries, in which users select emojis to reflect on their current emotional state. Emojis are pictographic symbols that represent facial expressions, objects, or abstract concepts, and help users express tone and intent in digital communication. Prior research has demonstrated that emojis can effectively convey emotions, moods, and physical states [35] In addition, passive data derives from smartphone sensors and include for Feelee the tracking of step count and sleep duration. By combining these self-reports emoji’s with passively collected data, Feelee provides a more comprehensive overview of adolescents’ daily emotional and behavioral experiences.

In forensic settings, tools like Feelee may provide valuable support in bridging the gap between therapeutic demands and adolescents’ limited emotion regulation capacities. Effective emotion regulation, as conceptualized in leading theoretical models like Gross [36,37] and Thompson [38,39] involves the ongoing process of monitoring, evaluating and modifying emotional experiences and responses. These processes correspond to three core skills: (1) emotional recognition, reflecting the initial stage of awareness and attentional deployment, (2) reflection, aligning with the appraisal phase in which individuals assign meaning to emotional experiences, and (3) emotional management, referring to the modulation of responses through the selection and application of regulatory strategies [40]. While research has mainly explored these skills as separate constructs, the evidence consistently highlights their individual contribution to emotional functioning. For example, training in emotion recognition has been associated with enhanced affective accuracy and improved behavioral outcomes [41,42]. Similarly, self-reflection has been linked to greater cognitive control and reduced emotional reactivity [43,44] and short-term interventions targeting emotion management have shown increased use of adaptive regulation strategies [45]. Despite this growing evidence base, these skills are rarely addressed as an interconnected, trainable set within interventions for adolescents.

Feelee responds to this need by translating these core components of emotion regulation skills into three practical steps: recognizing emotions through emoji’s, reflecting on emotional experiences, and offering tailored insights to support emotional self-management. An earlier pre-pilot study showed the feasibility and usability of Feelee within forensic outpatient setting [46]. Consistent with the pre-pilot, current Feelee usage comprises both independent app engagement and the incorporation of app-derived data into the therapeutic dialogue. For emotional recognition, adolescents use pre-selected emojis within the app to identify their current emotional states. In treatment sessions, discussing the meanings and nuances of these selected emotions fosters adolescents’ ability to recognize and articulate their feeling [47]. Similarly, reflection is supported through both in-app questions as well into the therapeutic discussions that explore the context of reported emotions. This process is further enhanced by Feelee’s weekly overviews, which combine the active self-reported emotional data with passively collected behavioral data (e.g., physical activity and sleep patterns). These overviews assist clinicians and adolescents in identifying emotional and behavioral patterns, facilitating collaborative reflection during therapy. Through this reflective process, adolescents gain greater insight into their emotional and behavioral functioning, which are essentials enhance effective emotion regulation [48–50]

Beyond emotion regulation, the integration of technologies like Feelee into treatment, offers new opportunities to better meet the needs and engagement of adolescents in forensic outpatient care [51]. Tools like Feelee better aligned with the way adolescents communicate digitally and their familiarity with using these tools to express experiences and emotions. The use of personalized, real-time data in a visual format is likely to fit better with their way of processing and expressing emotions than traditional, verbally demanding approaches [26,52]. Additionally, the continuous and objective nature of mobile data collection may help reduce stigma by providing a more impartial form of self-monitoring, in contrast to clinical ‘snapshot observations.’ This approach promotes a greater sense of being understood, making mental health support feel more accessible and less judgmental [53]. The use of Feelee in treatment also encourages shared goals, collaborative tasks, and joint decision-making, which positively contribute to the therapeutic alliance [52,54]. Outside of treatment sessions, Feelee lowers the threshold for adolescents to practice emotion regulation skills independently, thereby promoting self-management behaviors [35,55]. Altogether, Feelee may strengthen adolescents’ motivation and therapeutic relationships, ultimately leading to greater engagement and adherence to treatment.

### Study aims

The current study aimed to explore the initial effectiveness of the Feelee app in addition to treatment as usual for adolescents receiving forensic outpatient care. A multiple single-case experimental design (SCED) with both quantitative and qualitative components was applied across two research sites. The primary objective was to examine the effects of Feelee on three core components of emotion regulation targeted by the app: emotional recognition, reflection, and management, assessed through daily measurements. As a secondary objective, we examined changes in (a) emotional developmental factors, including emotional differentiation, awareness, self-reflection and self-insight, and (b) treatment-related factors, such as treatment motivation and therapeutic alliance. These secondary constructs were assessed at pre-, post-, and follow-up assessments. The third and final objective concerned the qualitative component, which explored both adolescents’ and clinicians’ experiences with the use and integration of Feelee in therapeutic practice. This qualitative component aimed to provide insight into their perspectives on the perceived usefulness and clinical relevance of Feelee in supporting emotion regulation, thereby informing further refinement and implementation.

## Methods

The protocol of this study was previously published [46]. Furthermore, the methodology and results have been reported in accordance with the Single-Case Reporting Guideline in Behavioral Interventions (SCRIBE) 2016 [56].

### Study design

In this study, a non-randomized multiple Single Case Experimental Design (SCED) across multiple participants was applied (Fig 1). A SCED involves repeated measurements at the individual level, comparing phases with and without the intervention across different phases [57]. This design is particularly suited for examining preliminary intervention effects in small, heterogeneous clinical populations that are often difficult to include in large-scale effectiveness studies such as randomized controlled trials. In the present study, neither randomization nor concurrent baselines across participants were implemented, as the length and timing of study phases varied naturally within routine clinical practice. Blinding of participants, clinicians, and researchers was not feasible given the nature of the design. Nevertheless, SCED provides a robust framework for an in-depth, initial exploration of the potential effects of Feelee among adolescents receiving forensic outpatient care [56].

**Fig 1.**
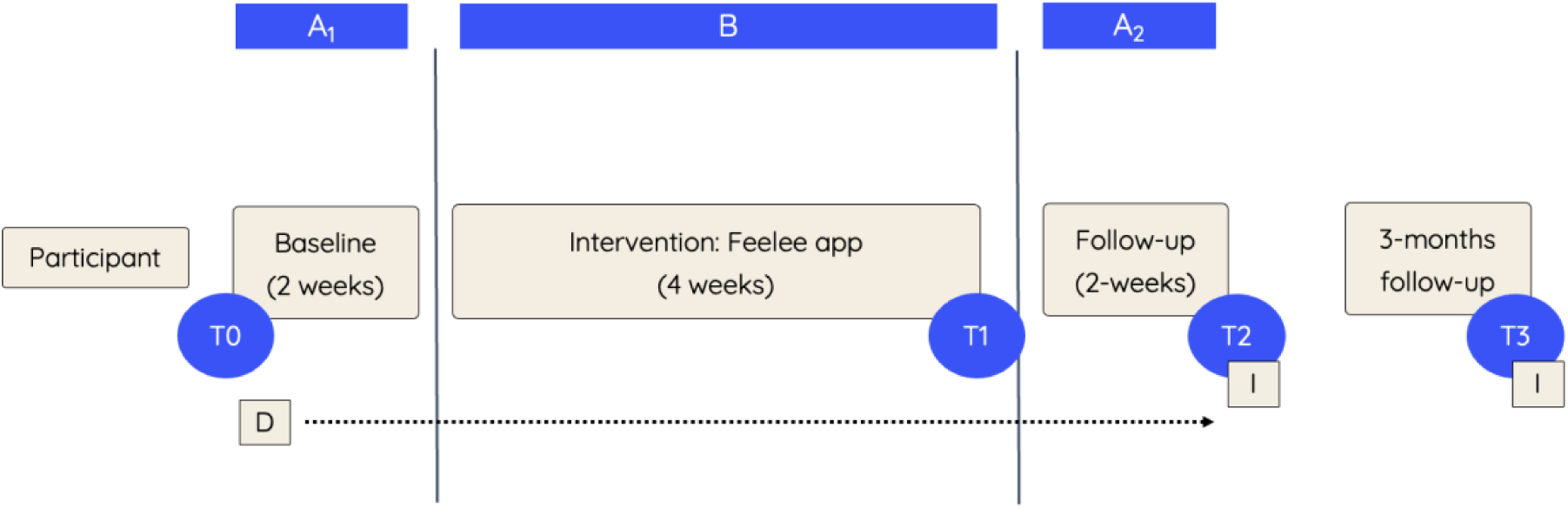
Overview of Single Case Experimental Design and Measurements. *Note*: A1: baseline; B: Intervention; A2: follow-up; T0: premeasurement; T1: postmeasurement; T2: follow-up measurement; T3: 3-months follow-up measurement; D: daily questionnaires; I: semi-structured interviews.

The SCED applied in the current study comprised a 2-week baseline (A_1_), a 4-week intervention (B), and a 2-week follow-up phase (A_2_). Throughout all phases, participants completed a daily questionnaire assessing various aspects of emotion regulation. In addition, pre- (T0), post- (T1), and follow-up (T2) assessments were conducted to facilitate a more comprehensive analysis of the potential mechanisms underlying effects of Feelee on emotion regulation. Three months after the initial follow-up measurement, a long-term follow-up assessment (T3) was conducted to evaluate the potential sustained effects of Feelee. Furthermore, semi-structured interviews were administered at T2 and T3 to gain deeper insight into participants’ perceived effectiveness of and experiences with Feelee as part of their treatment.

### Participants

Participants were recruited from outpatient forensic teams at two mental healthcare institutions in the Netherlands (Levvel and Inforsa) between 15 May 2023 and 15 October 2024. Potential participants were initially screened for eligibility by a research staff member in consultation with the responsible care provider(s). The inclusion criteria were as follows: (1) the participant was aged between 12 and 23, (2) was receiving individual and/or family-focused counseling or treatment at Levvel or Inforsa at the time of the study, (3) had an expected treatment duration of at least three additional months, (4) owned a personal smartphone with an iOS (iPhone) or Android operating system, and (5) possessed basic proficiency in smartphone use. Exclusion criteria included: (1) the presence of severe psychiatric conditions, such as psychosis or a high risk of suicide, (2) insufficient comprehension of spoken and written Dutch, or (3) lack of access to a personal smartphone with an iOS (iPhone) or Android operating system. In cases where eligibility was uncertain, a supervising senior clinician was consulted. Ultimately, 22 adolescents consented to participate. The overall dropout rate during the study was 36% (7 participants), which is relatively low given the forensic population [22,58]. The primary reasons for dropout included loss of interest in participating, prolonged disengagement from healthcare services (>3 months), or discontinuation of treatment due to external circumstances. An overview of the recruitment and participation process is presented in Fig 2.

**Fig 2.**
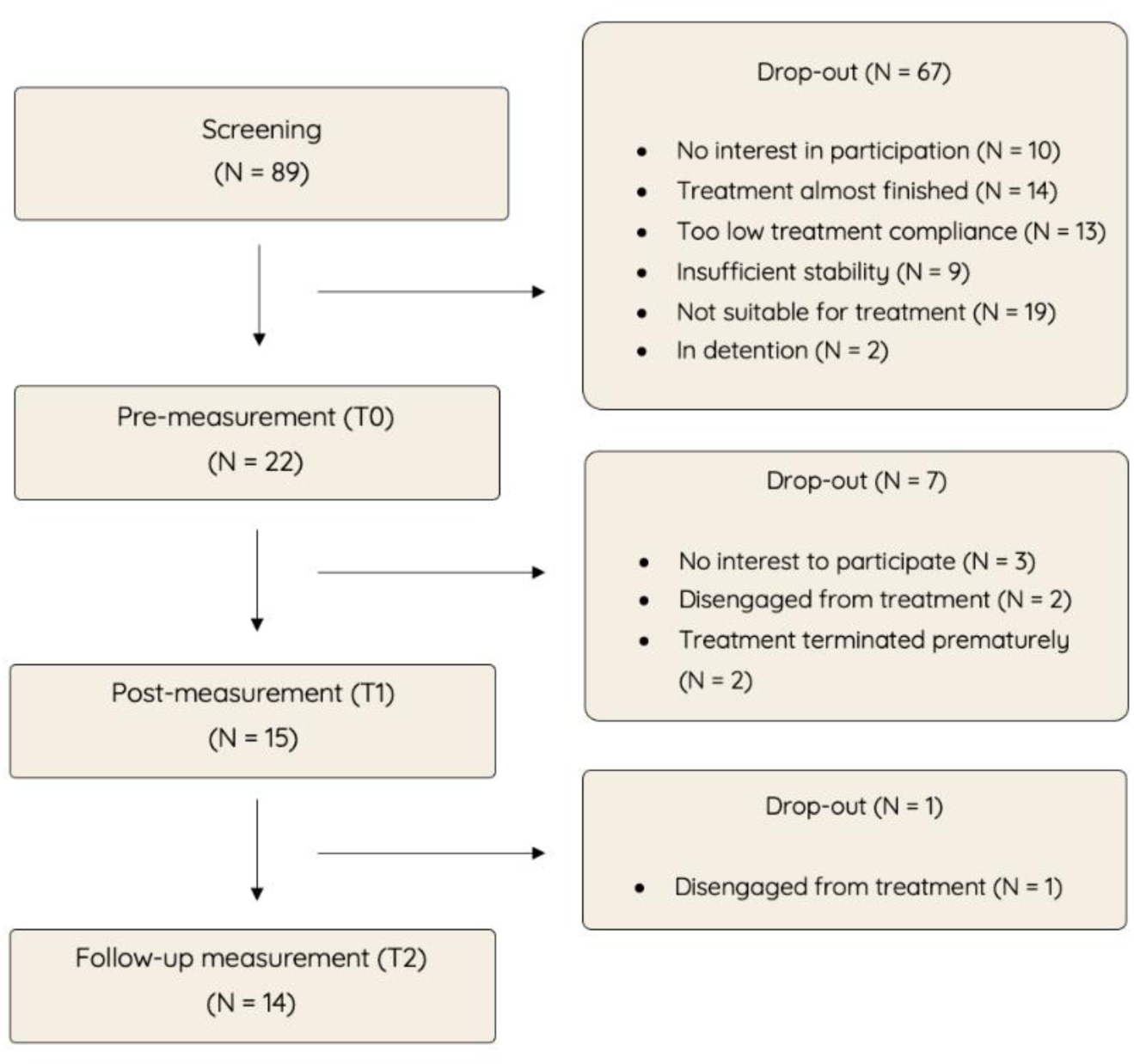
Recruitment and participation flow-chart.

### Procedure

At study onset, clinicians received a training on the study protocol and in- and exclusion criteria. Clinicians conducted the initial screening and eligibility, as they maintain regular contact with the adolescents. Eligible adolescents were briefly informed about the study and the Feelee app by their clinician. Those who expressed interest were scheduled for an appointment with a researcher, typically before or after a routine treatment session, during which detailed information about the study and the app was provided. If the adolescent remained interested, the consent form was reviewed, and pre-measurement (T0) appointment was scheduled, allowing a minimum of seven days for consideration. Written informed consent was obtained from the adolescent and, when applicable, from a parent or legal guardian for participants under 16 years of age.

Following informed consent, T0 was conducted, and the M-path app was installed to administer daily questionnaires. Participants received daily reminders to complete these questionnaires throughout the 2-week baseline phase, the 4-week intervention phase, and the 2-week follow-up phase. After completion of the baseline phase, participants commenced use of the Feelee app. Upon completion of the intervention phase, the T1 assessment was conducted, during which the Feelee app was uninstalled. The follow-up phase continued for two weeks with daily questionnaire completion, concluding with the T2 assessment and a semi-structured interview. Additionally, a final interview was conducted with the involved clinician. Three months following study completion, participants were re-contacted and invited to participate in a follow-up assessment (T3), which included questionnaire administration and a semi-structured interview. Participants received compensation for each completed questionnaire, including daily questionnaires and extended assessments (T0, T1, T2, and T3), as well as for each emoji entry made within the Feelee app. During T0, the researcher and the participant agreed on the frequency of updates regarding compensation.

### Measures and materials

In this section we first provide a more detailed description the Feelee app, the intervention tool being studied. Subsequently, we outline the primary outcome, emotion regulation, which was assessed through daily repeated measurements. This is followed by an overview of the secondary outcome measures, which were administered at pre-intervention, post-intervention, and follow-up. Finally, the third objective, exploring perceptions of participants and involved clinicians regarding the effectiveness and usage Feelee. This was addressed through semi-structured interviews, for which methodological details are described in the final part of this section. An overview of all measures and corresponding assessment time points is provided in Table 1.

**Table 1.** Measures, instruments and moment of assessment.

| Objectives | Instrument | T0 | DQ | Intervention | T1 | T2 | T3 |
| --- | --- | --- | --- | --- | --- | --- | --- |
| **Primary** |  |  |  |  |  |  |  |
| Emotion regulation | Selected items<br>DERS-18<br>RESS-EMA | x | x |  | x | x | x |
| **Secondary** |  |  |  |  |  |  |  |
| Emotional differentiation | PANAS | x |  |  | x | x |  |
| Emotional awareness | MAIA (subscale) | x |  |  | x | x |  |
| Self-insight and reflection | SRIS-Y | x |  |  | x | x |  |
| Treatment motivation | ATMQ | x |  |  | x | x |  |
| Treatment alliance | WAV-12 | x |  |  | x | x |  |
| **Other** |  |  |  |  |  |  |  |
| Demographic information | Questionnaire | x |  |  |  |  |  |
| Treatment integrity<sup>1</sup> | Questionnaire |  |  | x |  |  |  |
| Medical file information | File information | x |  |  |  |  |  |
| Evaluation Feelee app | Semi-structured<br>interview |  |  |  |  | x | x |

*Note:* T0: premeasurement; D: daily questionnaires; T1: postmeasurement; T2: follow-up measurement; T3: 3-months follow-up measurement.
<sup>1</sup> Completed by the clinician

#### Feelee App

At the start of the intervention phase, participants installed the Feelee app, (version 1.1.0), on their personal devices (iOS or Android). The Feelee app is owned by Garage2020, an innovation network within the youth care sector, and is free to use. In this study, participants received a pre-generated Feelee account linked to a non-personal email address to facilitate a smooth installation process and safeguard their privacy. During the use of Feelee, users are notified three times a day (by default at 8 a.m., 12 p.m., and 9 p.m., although these times can be adjusted) to select an emoji that reflects their current emotional state. This is followed by three brief reflective questions, ‘How come you feel this way?’, ‘What are you doing?’, and ‘Who are you with?’, which can be answered by selecting pictorial options or providing a written response (Fig 3). By combining these self-reports with passively collected data (e.g. sleep and number of steps), Feelee provides a more comprehensive picture of adolescents’ daily emotional and behavioral experiences. During the intervention phase, participants used Feelee independently by entering at least one emoji daily and permitting access to step and sleep data from their phone’s health app. Clinicians discussed these summaries during treatment sessions using a four-step approach of emotion regulation approach: (1) recognizing emotions via emojis and bodily experience, (2) reflecting on entry moments and context, (3) identifying emotion patterns linked to activities or social factors, and (4) discussing the influence of activity and sleep on emotions.

**Fig 3.**
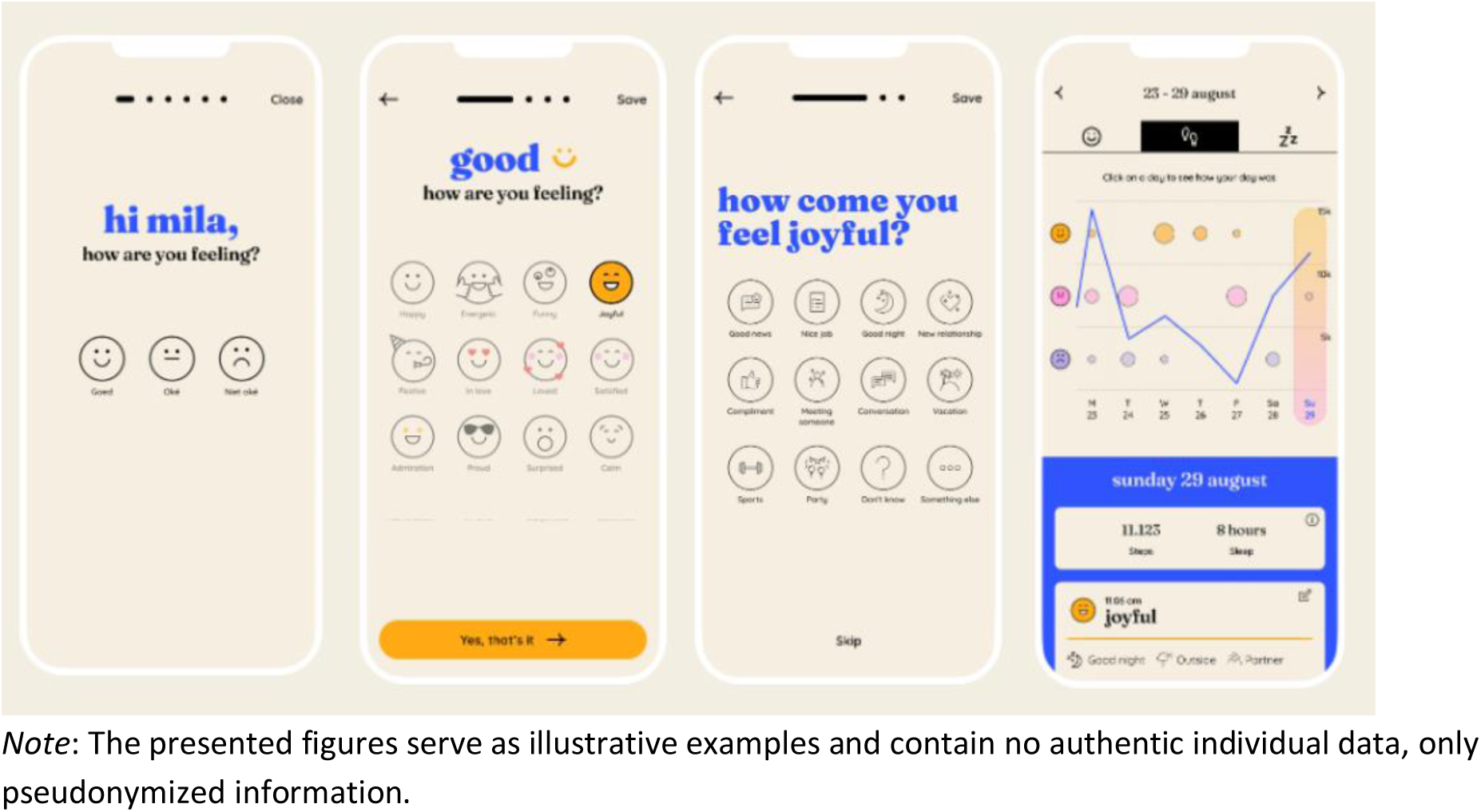
Screenshots of the Feelee app.

#### Emotion regulation

The primary outcome was assessed through daily questionnaires throughout the ABA design. To facilitate this, the M-path app was used to administer the daily questionnaire. Participants received a notification at a preferred time each day to complete the questionnaire, and if it was not completed, a reminder notification was sent after 120 minutes. To enable daily data collection, six items were selected to reflect the three prescribed phases of emotion regulation targeted by Feelee: recognition, reflection, and management. All items were adapted from the ‘Difficulties in Emotional Dysregulation Scale’ (DERS-36) [59] or ‘Regulation of Emotion Systems Survey for Daily Usage’ (RESS-EMA) [60]. For recognition, two items were included: recognition (clarity) and suppression. In this context, suppression was conceptualized as an automatic, unconscious response occurring early in the emotional process. For reflection, two compression items were included targeting rumination and emotional reappraisal. For management, impulse control and distraction were assessed. Responses could be provided on a 5-point scale, ranging from ‘strongly disagree’ to ‘strongly agree’ (DERS-36) [59] or scale from 0 = not at all to 100 = very much (RESS-EMA) [60]. For recognition (clarity) and manage (impulse) items, scores were recoded to ensure that higher scores consistently reflected higher levels of each construct.

Secondary objectives were assessed at pre-, post-, and follow-up time points to gain a more in-depth understanding of changes in (a) emotional developmental factors and (b) treatment-related factors, both at the individual and group level.

#### Emotional differentiation

Emotional differentiation was assessed using the Positive and Negative Affect Schedule (PANAS) [61] a 20-item scale that required participants to rate 10 positive and 10 negative emotions on a 5-point Likert scale ranging from ‘Not at all’ to ‘Extremely.’

***Emotional awareness*** was measured using the Multidimensional Assessment of Interoceptive Awareness (MAIA) [63]. In this study, only the ‘Emotional Awareness’ subscale was administered, which consisted of 32 items across 8 subscales rated on a 6-point scale from ‘Never’ to ‘Always.’ Treatment characteristics, including motivation and alliance, were also assessed.

#### Self-reflection and insight

To assess self-reflection and insight, the Self-Reflection and Insight Scale for Youth (SRIS-Y) [62] was utilized. This 17-item scale was rated on a 6-point Likert scale from ‘Strongly disagree’ to ‘Strongly agree,’ and included two subscales: Self-reflection and Insight, each providing separate scores.

**Treatment motivation** was measured using the Dutch Adolescent Treatment Motivation Questionnaire (ATMQ) [64], a self-report scale with 11 items rated on a 3-point Likert scale: ‘Not true,’ ‘Somewhat true,’ and ‘True.’

***Therapeutic alliance*** was assessed by the Working Alliance Inventory-12 (WAV-12) [65], a 12-item self-report questionnaire, was used. The WAV-12 evaluated the collaboration between the clinician and the adolescent on a 5-point Likert scale from ‘Rarely or never’ to ‘Always,’ with higher scores indicating a stronger therapeutic alliance.

***Treatment integrity*** was assessed using a self-developed questionnaire comprising three multiple-choice questions on whether a treatment session occurred that week, whether Feelee data were available, and whether the data were discussed. One open-ended question allowed for additional comments on the use of Feelee in treatment

The third and final objective aimed to gain an understanding of the use and experiences of adolescents and therapists with the Feelee app, assessed during both follow-up measurements.

#### Semi-structured interviews

The interviews focused on perceived effects on emotional skills and treatment-related factors, such as motivation and therapeutic alliance. Participants reflected on observed changes during therapy and the possible role of the app in these developments. Additional topics included the use of other smartphone apps and the integration of data into treatment. These interviews provided valuable insights to inform further development of the app and treatment approach.

### Data-analysis

The analysis of the primary objective was divided in two parts. First, we conducted visual inspection analyses on individual level using a *Shiny app* for Single-Case experimental Design [66]. For each participant, a graphical representation was generated to display the scores for each item of the daily emotion regulation questionnaire, connected over time. Participants needed at least five data points per phase to ensure reliable inspection [67]. The visual inspection focused on (a) the variability between data points, (b) changes between phases based on median scores, and (c) trends within and across phases. Group-level changes and trends were summarized using boxplots of item-level score distributions per phase, generated in R Statistical Software (version 4.2.1) [68] To enhance interpretability, the original 5-point Likert scale responses for the recognition (clarity) and management (impulse control) items were transformed to a 0–10 scale through linear rescaling. Additionally, randomization tests were conducted in Shiny apps as well [66]. Cohen’s d was calculated for each individual participant and averaged at group level, with effect sizes interpreted as 0.2 (small), 0.5 (medium), and 0.8 (large) [69]. Subsequently, a permutation test was performed [70]. This test does not correct for autocorrelation, as it is assumed that the scores on the six daily items do not exhibit substantial autoregression. Applying a correction for autocorrelation could therefore result in spurious effects. Tests at individual participant level were performed one-sided in the hypothesized direction. Individual *p*-values were obtained by repeatedly randomizing scores between phases and calculating the proportion of differences equal to or exceeding the observed. At group level, the sum of individual *p*-values was compared to the null distribution, using the properties of p-values that they are uniformly distributed between 0 and 1. Last, Tau-U scores were calculated to quantify change magnitude and direction per participant. Unlike the permutation test, Tau-U accounts for undesirable trends within phases, offering a more precise intervention effect estimate [71]. Tau-U values range from −1 to 1, with effect sizes interpreted as <0.2 (small), 0.2–0.6 (moderate), 0.6–0.8 (large), and >0.8 (very large) [72]. For all statistical tests a type 1 error rate of .05 was used.

Secondary outcome measures were analyzed using IBM SPSS (version 17) and R Statistical Software. For emotion differentiation, sum-scores were calculated separately for both positive affect (PA) and negative affect (NA) scales. Paired-sample t-test assessed changes in standard deviation scores for each subscale. For emotional awareness, self-reflection, insight, motivation, and therapeutic alliance, randomization tests compared mean scores between time points (T0, T1, T2) using *t*-statistics. The observed *t* was evaluated against a reference distribution from randomized data. The *p*-value reflected the proportion of more extreme randomized *t*-values. Individual changes were also assessed with the Reliable Change Index (RCI), calculated as the difference between time points divided by the standard error [73]. An RCI-score of ±1.96 indicates statistically reliable change, suggesting a meaningful change [74].

The semi-structured interviews were recorded and transcribed. One participant preferred not to be recorded. Therefore, a summary report of the interview was made. The transcripts and this report were independently coded by two researchers using MAXQDA (version 24). A thematic analysis was applied: several themes were pre-established based on the interview topic lists, and new themes were identified during the analysis of the transcripts. In case of uncertainties regarding themes or codes, the coders consulted each other, and if disagreements persisted, a third researcher was consulted [75].

### Ethical Considerations

This research project was approved in April 2023 by the independent Medical Ethics Committee of the Vrije Universiteit Medical Center (reference number: 2022.0398). As the study involves a clinical application of a smartphone as an integrated part of treatment, the Medical Device Regulation (MDR) applies. Therefore, the study was evaluated not only on ethical aspects but also on technical functionality and safety. Both the research data and the Feelee data were processed and stored in accordance with the General Data Protection Regulation (GDPR). Furthermore, together with the privacy officer of Amsterdam UMC, a Data Protection Impact Assessment (DPIA) and an evaluation of confidentiality, integrity, and availability (CIA) were conducted to identify potential risks and measures for data collection and storage.

## Results

The results are presented at the group level to provide a structured and comprehensive overview of the observed trends and patterns. Rather than presenting individual case outcomes in detail, the findings focus on overarching developments in the primary and secondary outcome measures. In addition, key themes from the semi-structured interviews are presented, capturing how adolescents and clinicians experienced the use of the app in clinical practice. More detailed results on individual cases can be found in S1 File. Furthermore, to illustrate the heterogeneity of individual trajectories, three different cases are presented in more detail in S1 File, S2 File, S3 File and S4 File. Each case represents a markedly different pattern of Feelee usage and variation in both timing and extent of observed effects, offering a more nuanced perspective on individual app usage and effectiveness in clinical practice. The names that are used in these individual cases are pseudonymized.

### Descriptive characteristics

The majority of 22 participants were male (86%), which is characteristic of the forensic population, where approximately 80% of adolescents in care are male (van der Laan et al., 2024; CBS Statline, 2024). Notably, all participants (100%) had parents who were either divorced or separated. Most participants (59.5%) resided permanently with one of their parents. Additionally, 68% of participants reported having been convicted by a court of law, while the remaining 32% had not been convicted but were receiving treatment as prevention. The participants were provided with various forms of therapeutic intervention, which are described in more detail in Table 2.

**Table 2.** Demographic characteristics.

| **Demographic characteristics** | **N = 22** |
| --- | --- |
| **Age (mean)** | 17,8 |
| **Gender (%)** |  |
| Male | 19 (86%) |
| Female | 3 (14%) |
| **Migration background** |  |
| First-generation migration background | 4 (18%) |
| Second generation migration background | 7 (32%) |
| Dutch background | 11 (50%) |
| **Relation status parents** |  |
| Separated or divorced | 22 (100%) |
| **Living situation** |  |
| Independently | 3 (14%) |
| Alternating between mother and father | 3 (14%) |
| With father or mother | 13 (59,5%) |
| With a relative | 1 (4,5%) |
| Supported housing | 2 (9%) |
| **Daily activities** |  |
| Non | 4 (18%) |
| School, specifically | 17 (77%) |
| • *Secondary school* | 4 (18%) |
| • *Vocational education* | 10 (45%) |
| • *University of applied sciences* | 1 (4,5%) |
| • *Other* | 2 (9%) |
| Employed | 1 (4,5%) |
| **Convicted by a judge** |  |
| Yes | 15 (68%) |
| No | 7 (32%) |
| **Treatment approach** |  |
| Family therapy combined with individual therapy | 5 (23%) |
| • *Relational Family Therapy (RFT)* | 2 (9%) |
| • *Systemic Family Therapy* | 3 (14%) |
| Individual Therapy | 15 (68%) |
| • *Aggression Regulation Therapy* | 2 (9%) |
| • *Cognitive Behavioral Therapy (CBT) (focus on enhancing emotion regulation skills)* | 5 (23%) |
| • *Offense analysis* | 3 (14%) |
| • *Social Support* | 3 (14%) |
| • *Trauma therapy (TF-CBT)* | 2 (9%) |
| Unknown | 2 (9%) |

### Primary outcome

First, cases were inspected on the presence of a sufficient number of data points (i.e., more than five) in each study phase. Among the participants who dropped out, one individual who discontinued participation during the follow-up phase had sufficient data points in the preceding phases and was therefore retained in the analyses. One participant who completed all phases was excluded due to insufficient number of data points. Consequently, 14 participants were included in the analyses, with completed data across all phases available for 13 participants. Furthermore, study progress and treatment integrity were evaluated based on clinician reports during the intervention phase. For one participant (7%), the app was discussed weekly. For 5 participants (36%), Feelee data were addressed in 2–3 of the 4 treatment sessions. For the remaining 7 participants (50%), the app was discussed once or not at all. No serious or adverse events occurred during the study. A detailed overview of treatment integrity is provided in S1 File.

To access changes in the emotion regulation across study phases, visual inspections and randomization tests were conducted. Fig 4 presents a boxplots of emotion regulation items for each phase, including both individual and group means scores. Table 3 shows the corresponding effect sizes, p-values, and the median of individual Tau-U scores for each phase comparison. Full results for all individual participants are provided in S1 File.

**Fig 4.**
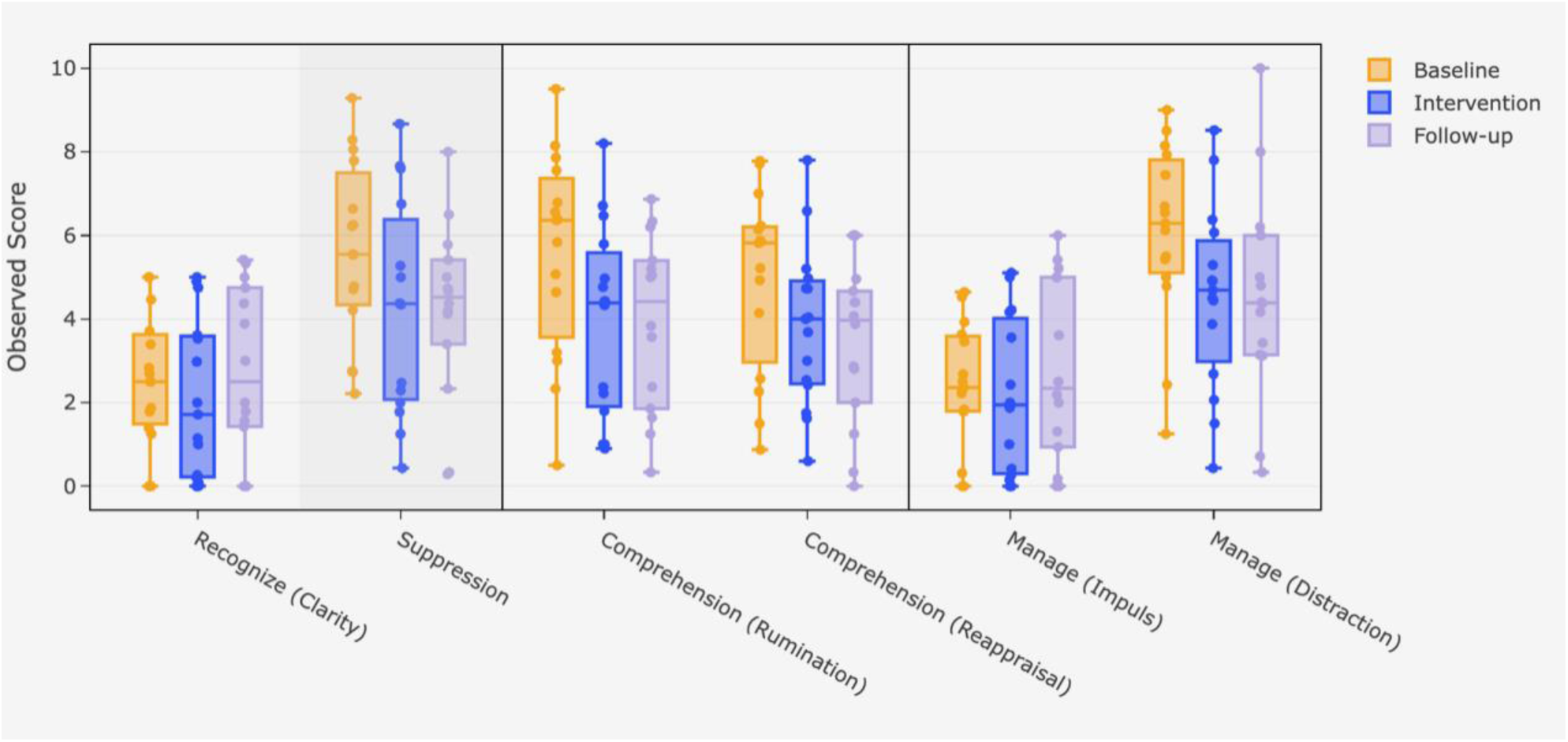
Boxplot for emotion regulation items for each phase.

**Table 3.** Effect sizes (Cohen’s d), p-values and number of significant cases from normal permutation test, and median individual TAU-U scores for each phase comparison.

|  | *N* | Expected direction | Cohen's *d* | *p*-value (median) | Significant Cases | Tau-U<sup>1</sup> (group) | Significant Tau-U Cases |
| --- | --- | --- | --- | --- | --- | --- | --- |
| **Recognize (Clarity)** |  |  |  |  |  |  |  |
| Baseline - intervention | 14 | (B<I) | -0.28 | 0.081 | 6 | -0.02 | 6 |
| Intervention - follow-up | 13 | (I<F) | 0.252 | 0.995 | 7 | 0.2 | 2 |
| Baseline - follow-up | 13 | (B<I) | -0.098 | 0.793 | 5 | 0.2 | 2 |
| **Suppression** |  |  |  |  |  |  |  |
| Baseline - intervention | 14 | (B>I) | 0.584 | 0.001* | 9 | -0.165 | 4 |
| Intervention - follow-up | 13 | (I>F) | 0.075 | 0.37 | 5 | 0.01 | 1 |
| Baseline - follow-up | 13 | (B>I) | 0.535 | 0.001* | 9 | -0.09 | 3 |
| **Comprehension (Rumination)** |  |  |  |  |  |  |  |
| Baseline - intervention | 14 | (B<I) | 0.693 | >.999 | 5 | -0.215 | 6 |
| Intervention - follow-up | 13 | (I<F) | 0.049 | 0.274 | 4 | -0.02 | 2 |
| Baseline - follow-up | 13 | (B<F) | 0.614 | >.999 | 8 | -0.25 | 5 |
| **Comprehension (Reappraisal)** |  |  |  |  |  |  |  |
| Baseline - intervention | 14 | (B<I) | 0.401 | >.999 | 7 | -0.06 | 5 |
| Intervention - follow-up | 13 | (I<F) | 0.293 | 0.741 | 3 | -0.19 | 2 |
| Baseline - follow-up | 13 | (B<F) | 0.401 | >.999 | 6 | -0.24 | 6 |
| **Manage (Impulse)** |  |  |  |  |  |  |  |
| Baseline - intervention | 14 | (B<I) | -0.107 | 0.529 | 6 | 0.095 | 4 |
| Intervention - follow-up | 13 | (I<F) | 0.361 | 0.913 | 3 | 0.3 | 4 |
| Baseline - follow-up | 13 | (B<I) | 0.057 | 0.823 | 4 | 0.16 | 4 |
| **Manage (Distraction)** |  |  |  |  |  |  |  |
| Baseline - intervention | 14 | (B<I) | 0.623 | >.999 | 4 | -0.24 | 6 |
| Intervention - follow-up | 13 | (I<F) | -0.017 | 0.864 | 4 | 0.1 | 1 |
| Baseline - follow-up | 13 | (B<F) | 0.644 | >.999 | 4 | -0.09 | 3 |

*Note.* Individual TAU-U scores are presented in S1 File.
\* Indicates $p < 0.05$
<sup>1</sup> TAU-U group-scores are based on median scores from total participant (N).

For the first step of emotion regulation, recognition (clarity), an increase was expected during intervention and follow-up compared to baseline. Visual inspection of the boxplot (Fig 4) showed a minimal decrease between baseline and intervention, followed by an increase during follow-up.

The randomization test, presented in Table 3, revealed a significant decrease only between the intervention and follow-up (*p* = .006), indicating a decline on recognition item during follow-up. The discrepancy between the increase in the boxplot (Fig 4) and the significant decrease in the randomization test (Table 3) is attributed to differences in the statistical procedures. In the boxplot, all observations are pooled within each phase, which means that participants with more observations disproportionately influence the group median. This can create an apparent increase in the median from the intervention to the follow-up phase. In contrast, the randomization test is conducted at the individual level, where each participant receives a p-value that is weighted equally in the group-level analysis. The significance of the group effect is then determined by the overall pattern of these equally weighted p-values, which may reveal an opposite effect to what is shown in the boxplot.

Besides the randomization test, Tau-U (Table 3) indicated small non-overlap effects between baseline and intervention for 6 participants, persisting in two during follow-up, suggesting significant change for 6 participants (43%) between baseline and intervention. On the second item, suppression, a decrease in scores was hypnotized and supported by Fig 4. Randomization test (Table 3) showed significant reductions from baseline to intervention and follow-up, indicating less suppression when participants entered the intervention phase. According to Tau-U scores, 4 participants noted a significant small non-overlapping effect between baseline and intervention. Effects between intervention and follow-up remained limited.

Regarding the reflection items, comprehension (rumination) and comprehension (reappraisal), an increase was expected starting at intervention and continuing during follow-up. Contrary to this, Fig 4 showed decreases during all phases. On both items, no significant differences were found between baseline and intervention or follow-up. However, reversed tests indicated significant reductions between baseline and intervention (*p* = .001) and between baseline and follow-up (p = .001), indicating participants reported lower engagement on both reflection items. Tau-U scores showed strong non-overlap effects for 3 participants (21%) on comprehension (rumination) and small effects for comprehension (reappraisal) in 5 participants (36%) between baseline and intervention, with limited follow-up continuation.

For impulse (manage item), an increase was expected throughout study phases. Both Fig 4 and Table 3 confirmed this pattern. However, randomization test revealed no significant differences between phases. Tau-U scores did reveal a small but significant non-overlap effect between baseline and intervention for 4 participants (29%), persisting for 1 participant (7%) during follow-up. For distraction (manage item), an increase was expected starting in the intervention phase, continuing during follow-up. Instead, Fig 4 shows a notable decrease between baseline and intervention, followed by a slight increase at follow-up. Randomization test confirmed no significant effects in the hypothesized direction, but reversed tests showed significant decreases from baseline to intervention (*p* = .001) and follow-up (*p* = .001), indicating that participants noted less distraction throughout study. Tau-U scores showed small non-overlap effect for 6 participants (43%) with no further significant effects during follow-up.

### Secondary outcomes

We evaluated group differences on both (a) emotional development and (b) treatment factors across pre-, post-, and follow-up measurements.

#### Emotional developmental factors

Starting with emotional differentiation, we aimed to investigate whether there was an increase in variability in responses related to positive and negative affect. To assess this, we performed a paired-sample t-test based on the standard deviation (SD) to explore an increase in emotional variability over time. As shown in Table 4, the variability in positive affect did not significantly increase between T0 and T1, but a significant increase was shown between T1 and T2. In contrast, the variability in negative affect remained relatively stable across all three time points, with no significant differences observed between any of the measurement moments.

**Table 4.**
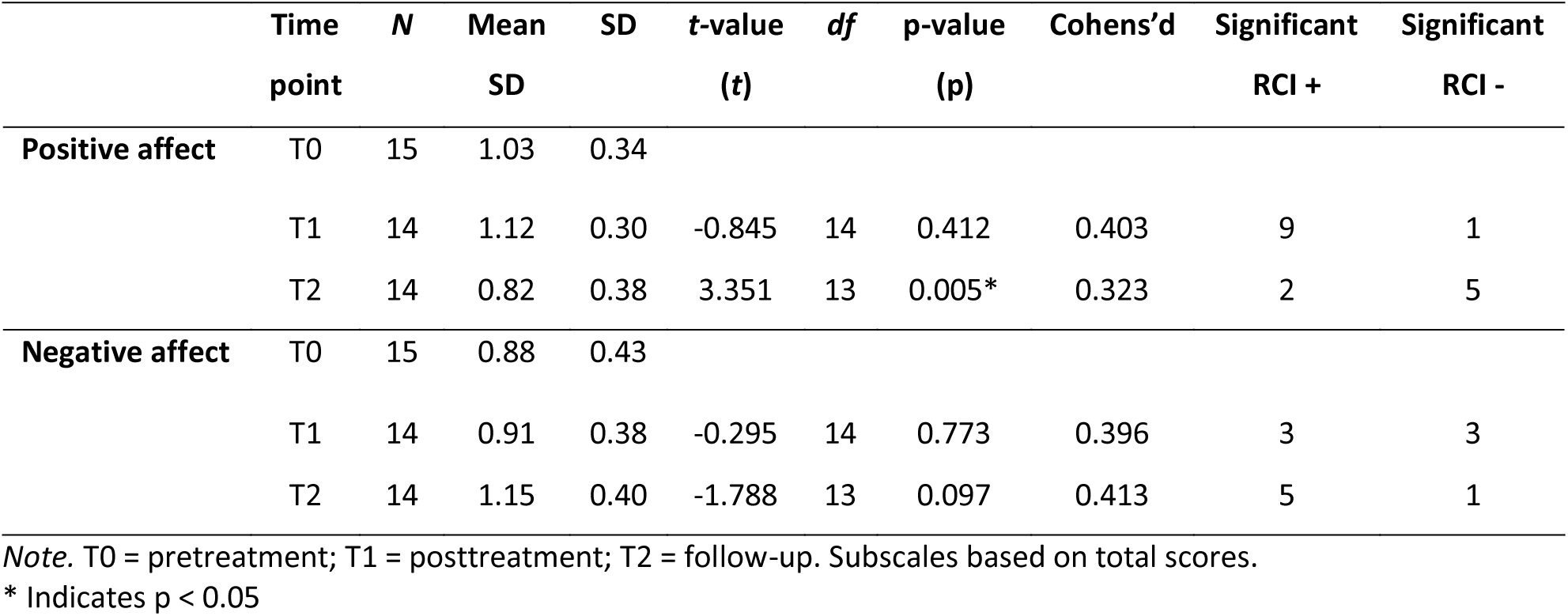
Changes in emotional differentiation across time points: Within-person SD scores and paired-sample t-test results for positive and negative affect.

|  | Time point | N | Mean SD | SD | t-value (t) | df | p-value (p) | Cohens'd | Significant RCI + | Significant RCI - |
| --- | --- | --- | --- | --- | --- | --- | --- | --- | --- | --- |
| Positive affect | T0 | 15 | 1.03 | 0.34 |  |  |  |  |  |  |
|  | T1 | 14 | 1.12 | 0.30 | -0.845 | 14 | 0.412 | 0.403 | 9 | 1 |
|  | T2 | 14 | 0.82 | 0.38 | 3.351 | 13 | 0.005* | 0.323 | 2 | 5 |
| Negative affect | T0 | 15 | 0.88 | 0.43 |  |  |  |  |  |  |
|  | T1 | 14 | 0.91 | 0.38 | -0.295 | 14 | 0.773 | 0.396 | 3 | 3 |
|  | T2 | 14 | 1.15 | 0.40 | -1.788 | 13 | 0.097 | 0.413 | 5 | 1 |

Note. T0 = pretreatment; T1 = posttreatment; T2 = follow-up. Subscales based on total scores.
\* Indicates $p < 0.05$

Next, we performed randomization tests for emotional awareness and both self-reflection insight subscales. As shown in Table 5, no significant changes were found for emotional awareness. RCI-scores showed a reliable increase in 1 participant between T1 and T2, no change between T0 and T2, and a reliable decrease in 1 participant between T0 and T1. For self-reflection, no significant increases were observed between T0 and T1 or T0 and T2, however, a significant increase occurred between T1 and T2. Reliable Change Index (RCI) scores indicated that 8 participants showed a reliable increase between T0 and T1, while 4 showed a reliable decrease; this pattern was similar across T0–T2 and T1–T2. Lastly, for insight, randomization tests showed no significant changes at any time point. RCI-scores revealed reliable increases for 3 participants between T0 and T1, 4 between T0 and T2, and 5 between T1 and T2, with similar numbers showing reliable decreases.

**Table 5.** Results of randomization test and significant Reliable Change Index (RCI) cases for secondary emotional outcomes.

|  | Time point | N | Mean | SD | p-value (p) | Cohens'd | Significant RCI + | Significant RCI - |
| --- | --- | --- | --- | --- | --- | --- | --- | --- |
| **Emotional awareness** | T0-T1 | 15 | 16.32 | 5.31 | 0.282 | 0.23 | 0 | 1 |
|  | T1-T2 | 14 | 18.07 | 5.43 | 0.047* | -0.51 | 1 | 0 |
|  | T0-T2 | 14 | 14.87 | 7.95 | 0.118 | -0.35 | 0 | 0 |
| **Self-reflection** | T0-T1 | 15 | 41.05 | 6.39 | 0.325 | 0.02 | 8 | 4 |
|  | T1-T2 | 14 | 43.57 | 8.01 | 0.044* | -0.51 | 8 | 3 |
|  | T0-T2 | 14 | 40.13 | 6.45 | 0.067 | -0.44 | 8 | 2 |
| **Insight** | T0-T1 | 15 | 23.64 | 6.28 | 0.328 | 0.1 | 3 | 3 |
|  | T1-T2 | 14 | 22.86 | 3.61 | 0.102 | -0.37 | 5 | 4 |
|  | T0-T2 | 14 | 22.13 | 5.54 | 0.218 | -0.22 | 4 | 3 |

*Note.* T0 = pretreatment; T1 = posttreatment; T2 = follow-up. Self-reflection and Insight were measured using sum-scores. Emotional awareness is based on the subscale mean score.
\* Indicates $p < 0.05$ .

#### Treatment factors

Randomization tests assessed also changes in treatment motivation and treatment alliance across the T0, T1, and T2 time points. As shown in Table 6, no significant changes were found in treatment motivation over time. Additionally, the Reliable Change Index (RCI) analyses did not indicate any reliable improvements. In contrast, treatment alliance showed a significant increase between T0 and T2. RCI-scores revealed a reliable improvement for 3 participants from T0 to T1, 2 participants from T0 to T2, and 3 participants from T1 to T2. Conversely, a slightly higher number of participants showed a reliable decrease: 4 between T0 and T1, 5 between T0 and T2, and 2 between T1 and T2.

**Table 6.** Results of randomization test and significant Reliable Change Index (RCI) cases for secondary treatment outcomes.

|  | Time point | N | Mean | SD | p-value (p) | Cohens'd | Significant RCI + | Significant RCI - |
| --- | --- | --- | --- | --- | --- | --- | --- | --- |
| Treatment motivation | T0-T1 | 15 | 24.95 | 3.15 | 0.112 | -0.42 | 0 | 0 |
|  | T1-T2 | 14 | 27.14 | 4.66 | 0.512 | 0 | 0 | 0 |
|  | T0-T2 | 14 | 26.80 | 3.49 | 0.17 | -0.26 | 0 | 0 |
| Treatment alliance | T0-T1 | 15 | 48.50 | 8.97 | 0.067 | 0.4 | 3 | 4 |
|  | T1-T2 | 14 | 47.57 | 10.53 | 0.354 | -0.1 | 3 | 2 |
|  | T0-T2 | 14 | 45.80 | 11.87 | 0.034* | 0.55 | 2 | 5 |

Note: T0 = pretreatment; T1 = posttreatment; T2 = follow-up. Treatment motivation was calculated using the mean score; treatment alliance is based on the sum-score. \* Indicates $p < 0.05$ .

### Qualitative results

Qualitatively, the result focus on perspectives of participants and clinicians regarding the Feelee experienced effectiveness and working elements in treatment. An overview of the themes and responses from both participants and clinicians are presented in Table 7.

**Table 7.** Overview of statements from both adolescents and clinicians semi-structured interviews.

| Theme | Adolescents (n = 12) | Clinicians (n = 11) |
| --- | --- | --- |
| **Users’ general experiences with Feelee** | Positive, helpful in treatment (n=10) | Positive, easy to use (n=4) and suitable to integrate in treatment (n=2) |
|  | More critical, forget to use it (n=1), app seen as childish (n=1) | More critical, limited engagement possibilities (n=2) and passive data issues (n=5). |
| **Effects of Feelee on emotion regulation** | To think more deeply about emotions (n=7) | Increased emotional awareness (n=4) and self-insight (n=3) |
|  | To pause and process emotions (n=2) | Increased emotional recognition (n=3) |
|  | Emotional release (n=2) | To think more about positive emotions (n=2) |
|  | No effects experienced (n=2) | No effects for emotional differentiation (n=2) |
| **Working elements of Feelee in treatment** | Recall emotions and situations (n=10) | Insight in daily emotions and activities outside treatment sessions (n=7) |
|  | Expressing emotions by emoji's verbally (n= 3) | Expressing emotions by emoji's verbally (n= 5) |
|  |  | Supportive to conversation in treatment (n = 9) |

First, regarding experiences of using the Feelee app, most participants reported being enthusiastic (n= 10). As one adolescent described: “*It was really easy, it actually blended quite well into my routine. (…) How I was feeling, or if something was bothering me, I could easily put it in.” (P19).* Two participants were more critical, citing forgetfulness (n=1) or perceiving the app as childish (n= 1): *“I always felt… I think it’s a bit (…) kind of childish or something.” (P5)*. Clinicians were also positive. Several found the app easy to use (n= 4) and suitable for integrating into treatment (n= 2), particularly to support emotional reflection: “*I think the Feelee app is a really easy and accessible way to talk about things, without it always feeling like an extra topic that had to be brought up during the individual sessions*.” (C7). Criticism focused mainly on limited engagement possibilities (n= 2) and issues with passive data (n= 5). One clinician noted a quick decline in use due to a lack of interaction: “*The core of the Feelee app is solid (…), but a next step would be being able to view more personalized input over time*.” (C2). Another clinician added: “*It’s frustrating when it doesn’t work properly, because it could really help you understand things better*.” (C8), referring to missing data on step count and sleep.

In terms of experiences regarding the effectiveness of Feelee, did a small majority of participants (n= 7) indicated that Feelee helped them to reflect more deeply on their emotions. As one participant described: *“I just started thinking about it more, like: ‘Oh, yesterday I felt that way, you know?’ I was kind of, uh, low on energy, for example or I was angry or something.”* (P9). Clinicians emphasized that this reflection was more about increased ‘awareness’ (n= 4) and self-insight (n= 3). One clinician noted: *“I actually noticed that when she started using the app, she became much more aware of what was happening within herself on a daily basis. She also began to think, ‘Hey, where is this coming from?”* (C1). Some adolescents (n= 2) mentioned that Feelee helped them pause and process emotions, while others (n=2) described it as a tool for emotional release. As one participant explained: *“It’s easy for me to put my emotions into an app like that, to remember how I felt on a certain day (…). And if, for example, I can’t talk to anyone about it, at least I still have the app.”* (P19). While adolescents noted these benefits, clinicians observed an increase in emotion recognition (n=3): “*It may sound simple, but with him, a lot of things just pass by. (…) It starts with recognizing and acknowledging emotions. I think for this adolescent, just being aware of that is already a big step forward.”* (C8). Clinicians also mentioned that Feelee sometimes supported reflection on positive emotions. However, a few participants (n=2) reported no noticeable emotional effects from using the app. According to clinicians, emotional differentiation remained limited. As one explains: “*It’s noticeable that this group is actually quite undifferentiated in their emotions, and that it was still difficult for them to do.”* (C7).

Concerning the working elements of Feelee in treatment, participants (n= 10) stated that the app’s weekly overview helped them recall emotions and past situations. Clinicians found this valuable for gaining insight into daily emotions and activities outside of treatment sessions. One clinician explained: *“I’m there, yeah, maybe one hour a week or sometimes two. But what someone is doing outside of that, well, you start to get more and more insight into that through an app like Feelee.”* (C6). Both participants (n=3) and clinicians (n=5) highlighted the usefulness of emojis for expressing emotions verbally during treatment. As one participant shared: *“(…) Sometimes I find it hard to explain things. So being able to show it like that in the figure and then reflect on it. That was really helpful.”* (P8). Similarly, a clinician noted: (…) “*When I asked questions like ‘Did you see your girlfriend?’, he couldn’t really express that. But with the Feelee app, (…) he could link it to an emoticon and tell a whole story from beginning to end*.” (C9). Additionally, clinicians found Feelee supportive for monitoring and discussing emotions and related behaviors in treatment. As one clinician said: “It did work in the sense of: “*Oh right, I need to keep an eye on that.’ So, I did ask about it. And that was really thanks to the Feelee app that made more alert to it.”* (C6).

## Discussion

The present study aimed to explore the initial effectiveness of Feelee in enhancing emotion regulation skills among adolescents in forensic outpatient care. Feelee was incorporated during a 4-week intervention phase, in addition to treatment as usual. The primary objective focused on improvements in three core components of emotion regulation: recognition, reflection, and management. Secondary objectives included a more in-depth examination of changes in (a) emotional developmental factors and (b) treatment-related factors, assessed at pre-, post-, and follow-up. And the third and final objective involved a qualitative exploration of both adolescents’ and clinicians’ perspectives on the effectiveness and use of the Feelee app.

Findings on the primary outcome showed a significant reduction in emotional suppression. However, contrary to expectations, decreases were also observed in items related to reflection and management (distraction). For the secondary outcomes, longer-term improvements emerged in positive emotion differentiation, emotional awareness and self-reflection. No clear change was observed in treatment motivation, while therapeutic alliance showed a significant increase over time. Qualitative data confirmed that both participants and clinicians were generally enthusiastic about the use of Feelee in treatment, although they also identified areas for improvement, such as technical challenges and limited integration of app data into therapy sessions. While results are presented at the group level, three illustrative case studies, included in the supplementary materials, offer a more detailed understanding of the heterogeneity in how Feelee was used and experienced across individual participants.

The results on the primarily outcome, emotion regulation, revealed interesting patterns for further discussion. First, a significant reduction in the suppression item indicates that participants suppressed their emotions less during and after using Feelee. However, no further improvements in the expected direction were observed for the other emotion regulation items: recognition, reflection, and managing. Supported by the qualitative results, does it suggest that Feelee is helpful in reducing suppression and support participants to engage in emotions. This can be considered as first important step before advancing to more complex strategies that rely on reflective and cognitive abilities [76,77]. This is in line with the growing literature emphasizing the critical role of targeting earlier stages of emotions regulation like recognition and (interoceptive) awareness in preventing disruptive behavior [19,78,79]. Moreover, these study findings build on a previous intervention study by ter Harmsel and colleagues in 2023, who observed increased interoceptive awareness following the use of a biofeedback app to detect early emotional signals such as stress or anger [80]. Together, these findings add to a growing body of evidence suggesting that enhancing emotion regulation should not be viewed entirely through the lens of foundational emotion regulation models like Gross [37,40]and Thompson [38]. While these models focus on selecting and implementing effective regulation strategies, assuming a baseline capacity for emotional recognition and cognitive reflection, they tend to overlook the importance of the initial stages of becoming aware of and identifying emotional states. Our results indicate that these early phases may constitute a critical target for intervention and hold meaningful potential for improving outcomes among adolescents in forensic outpatient settings.

Our findings on reduced suppression might also explain the reversed findings on distraction (managing). We initially hypothesized that distraction behaviors would increase due Feelee promotion of physical activity and sleep as activities designed to elicit positive emotions. In line with the reduction in suppression, it is possible that adolescents became more open to experiencing their emotions directly, reducing the need to rely on distraction. Therefore, distraction may have shifted in form and function: from maladaptive strategies aimed at avoiding emotional experiences, to more adaptive, goal-directed behaviors [39]. As these more constructive forms of distraction became integrated into daily routines, adolescents may no longer report distraction in a maladaptive sense, which may explain the decline observed during the use of Feelee.

Another noteworthy pattern in the primary outcome emerged for both reflection items: rumination and reappraisal. While we initially expected that Feelee would enhance participants’ reflection, the reversed analysis showed a significant decrease in self-reported rumination and reappraisal after the start of the intervention. This finding raises several considerations. It may be linked to the emotional capacities of adolescents in forensic care [19,21]. Participants primarily showed improvements in reducing suppression, an initial stage of emotion regulation. If adolescents are unfamiliar with the concept or practice of reflection, they may find it difficult to recognize or report progress in this area. Another explanation can be found in the treatment integrity data that revealed in the 4-week intervention that for over 50% of participants, the Feelee data was discussed once or not at all. This limited discussion could have hindered the training of reflection skills during the intervention, which potentially explaining participants’ lower self-reported reflection on the primary outcome. Last, despite pre-testing the daily questionnaire, it remains possible that both reflection items were not measured accurately, potentially introducing biases or leading to misinterpretations. For instance, the item ‘rumination’ can be interpreted as a reflection on a negative experience instead of a reflection of a potential positive experience. Therefore, a decrease could also indicate less reflection on negative experiences but potentially increase of positive experiences. Similarly, for reappraisal, participants may have interpreted ‘the situation’ in varying ways. These differences may have influenced which types of situations participants reflected on, thereby affecting the consistency and interpretability of their responses. It is plausible that these varying interpretations may have influenced the outcomes.

Beyond the primary outcomes, the secondary measures revealed several noteworthy effects, particularly at follow-up measurements. Specifically, improvements in positive emotional differentiation, self-reflection, and emotional awareness suggest a delayed yet meaningful development of emotional competencies among participants. Rather than showing immediate effects, the results suggest that Feelee may have initiated changes in these domains during the intervention phase, while actual improvements apparent at a later stage. This aligns with meta-cognitive research suggesting that these are a higher-order emotional skill, requiring cognitive and meta-reflective capacities that may not develop within a short 4-week intervention period, but may instead require continued engagement and repeated practice [81,82].

With regard to the secondary treatment-related factors, did we not find further improvement in participants’ motivation throughout the study. Notably, the high motivation scores at baseline indicate that the adolescents who participated were already motivated at the start of the study. However, this also highlights the complexity of reaching adolescents who are not motivated for treatment, as this group was not targeted in this study [83,84]. For treatment alliance, we first observed a slight decrease, followed by a significant improvement in participants’ reported alliance. These findings support earlier studies on the integration of digital elements in treatment, which have shown that such tools contribute to shared goal setting and foster a more shared language in treatment [85]. For adolescents, this enhances feelings of being understood and, conversely, helps clinicians better understand their patients and provide more tailored care [86]. Therefore, Feelee appears to be a promising tool for these adolescents to strengthen the therapeutic alliance and potentially increase their engagement in treatment.

### Strengths and limitations

This study has various strengths and limitation. A major strength is that this study belongs to a limited body of research investigating the initial effectiveness of a data-driven smartphone app aimed at enhancing emotional regulation skills among adolescents in forensic outpatient settings. By conducting this study, we contribute to addressing the lack of research on new technologies for adolescents in forensic outpatient settings. Second, although the forensic population is considered complex for research due to typically low engagement and high dropout rates, we were able to recruit and include a sufficient number of participants for the data analyses. Another strength is the use of a mixed-methods design, which enhanced the understanding of study outcomes and their implications for clinical practice. Finally, in addition to the study design, the research was conducted under real-world, uncontrolled circumstances, making the outcomes directly valuable for clinical implementation.

Nonetheless, our study has also several limitations. First, although the study design (SCED) incorporated structured baseline, intervention, and follow-up phases, the small overall sample size restricts the generalizability of the findings and calls for caution when drawing conclusions about the app’s effectiveness for a broader population. Nevertheless, SCEDs offer particular methodological advantages over traditional randomized controlled trials (RCTs), especially in exploratory research. They allow for detailed insights into individual change processes and contextual factors that often remain unexamined in standard group designs. Second, although recruitment efforts were as broad as possible, it was predominantly highly motivated adolescents who agreed to participate, or who were selected for participation by their caregivers. This self-selection may have introduced bias and could result in an overestimation of the intervention’s feasibility or perceived usefulness in the broader forensic population. Third, while Feelee collects various types of data, its integration into treatment remained limited. In practice, only the self-reported emoji-based mood data were actively discussed during therapy sessions. Additionally, technical issues hindered the passive data collection (e.g., activity or sleep metrics) in several cases. For these participants, the intervention was limited to its active components, reducing the scope of its originally intended functionality. Consequently, the digital phenotyping component for these individual cases was based solely on active data. Finally, the daily repetition of the daily questionnaire may have affected the validity of responses. The high frequency and repetitive nature of the items may have led some participants to respond in a socially desirable manner or to answer quickly without fully considering the content of each question. Such tendencies could reduce the accuracy of the data and reflect decreased engagement over time, especially in a population that may have limited tolerance for structured self-monitoring procedures [87].

### Implications for research and clinical practice

The findings of this study highlight the importance of focusing on earlier phases of emotion regulation, such as reducing emotional suppression could foster greater engagement with emotions, especially in forensic youth populations. While many interventions tend to emphasize managing emotions, our results suggest that the ability to fully experience emotions, with less avoidance, should be regarded as a valuable therapeutic goal in itself. Additionally, the findings indicate that more advanced emotion regulation skills, which require higher cognitive engagement, may develop after a longer period of usage. Therefore, Feelee shows promise as a tool that, when meaningfully integrated and actively discussed within treatment, can support the development of these fundamental emotion regulation skills.

In future research, larger-scale studies are needed to build on these initial findings, with particular focus on whether more advanced emotion regulation skills develop after prolonged use. Moreover, it is important to consider how research designs can be made more inclusive, particularly in forensic populations, where adolescents are often less willing to participate in research. Although low motivation should not be seen as a target in itself, this subgroup is frequently underrepresented in studies, while they may benefit substantially from low-threshold, digitally supported interventions. Future efforts should therefore explore how research procedures and tools can be adapted to better engage these adolescents, for instance by simplifying participation and increasing the accessibility and relevance of research activities. Another promising direction is to further investigate the use of Feelee as a screening tool. The Feelee data provide clinicians with valuable insights into the emotional functioning of adolescents, also outside treatment sessions, and help guide decisions regarding the focus and intensity of treatment. Especially in the early stages of treatment, Feelee may support adolescents in avoiding less and engaging more with their emotions before moving on to more advanced emotion regulation skills.

For clinical practice, our findings emphasize the importance of integrating app data into the therapeutic dialogue. Adolescents reported that reviewing their Feelee data during therapy helped them better understand their emotional patterns. However, treatment integrity showed limited implementation of discussing Feelee data within treatment. Therefore, clinicians are encouraged to actively incorporate these data into sessions, rather than treating the app as a stand-alone tool. This also highlights the need for further clinical guidance on the integration of a data-driven smartphone app, including how to effectively discuss and apply the Feelee data in treatment.

## Conclusion

This study showed that Feelee is a valuable tool in supporting early emotion regulation, particularly by reducing emotional suppression. These results suggest that Feelee may support early stages of emotion regulation by helping reduce suppression, a critical first step for this population. While no immediate effects were observed on reflection or management during the short intervention period, longer-term improvements in emotional differentiation, self-reflection and emotional awareness emerged. This initial decline may reflect increased emotional openness rather than poorer regulation. Regarding treatment outcomes, participant motivation remained stable, whereas the therapeutic alliance significantly improved. Qualitative findings supported these quantitative results, while also indicating that technical issues and limited integration of Feelee data into treatment discussions may have constrained the app’s full potential. The heterogeneity in outcomes across the three cases further underscores that the effectiveness and functioning of Feelee may differ depending on individual needs and context. Future research should examine whether prolonged use of Feelee can foster the development of higher-order emotion regulation skills, such as self-reflection and active emotion management. To support this, it is essential to involve both adolescents and clinicians in the design and evaluation process, in order to reach a broader population and to develop clear guidance on how to meaningfully integrate and discuss Feelee data within treatment.

## Data Availability

All relevant data are within the manuscript and its Supporting Information files.

## Supporting information

S1 File. Detailed results outcomes all participants

S2 File. Detailed result outcomes of case 1

S3 File. Detailed result outcomes of case 2

S4 File. Detailed result outcomes of case 3

## Acknowledgments

The authors would like to thank all adolescents and clinicians who participated in this study. We are also grateful to the clinicians who were not directly involved in the study but contributed to the recruitment process. Furthermore, we thank Mignonne Curiel, Aaliyah Hansen, Anne Kok, and Imara Semeijn for their valuable support during the preparation, recruitment, data collection, and data analysis phases of the study.

## Author Contributions

ML, LD, and AP were responsible for the conceptualization of the study and the development of the study design. ML coordinated the data collection and processing, wrote the manuscript, and performed the quantitative data analysis in collaboration with SB. ML and IS conducted the qualitative analysis. TP and RB provided operational support during recruitment and data collection. LD, SB, TP, RB and AP critically revised the manuscript. All authors read and approved the final version of the manuscript.

